# Cascading Feature Refinement with Channel-Wise Attention for Super-Resolution of Gastrointestinal Capsule Endoscopy

**DOI:** 10.64898/2026.11.25.26368968

**Authors:** Grace Katuri, Atchyuth Kumar Pairdha

## Abstract

Wireless capsule endoscopy (WCE) now enables endoscopic examination of the whole small bowel, albeit typically in a non-invasive, more comfortable, and generally painless manner. The physical limitations of such endoscopy technology (specifically, sensor array size, battery, and light constraints) usually dictate relatively low-resolution (LR) output video frames. Consequently, capturing, and thus detecting, fine or non-protruding gastroin-testinal (GI) abnormalities (e.g., small mucosal vascular lesions, mild angioectasias, and early mucosal neoplastic conditions) can sometimes be clinically sub-optimal. This paper presents a novel cascading feature refinement network involving Dense Shortcut Connections and Channel-wise Attention Mechanism (CFR-CA). The network operates on only the Luminance (Y) component of YCbCr colour space of the WCE images, exploiting high-frequency structures that are key to the GI diagnosis, while efficiently skipping processing steps on Chrominance (Cb, Cr) channels. We collected a dataset of 10,000 WCE frames from different institutions, pre-processing them by manually removing the frame corners to create our training dataset. We utilised bicubic down-sampling (× 4) to generate our LR and high-resolution (HR) pairs. Experimental results show that CFR-CA achieves the best RGB Peak Signal-to-Noise Ratio (PSNR), the best Structural Similarity Index Measure (SSIM) on both the Y and RGB channels, and the best Learned Perceptual Image Patch Similarity (LPIPS) among contemporary techniques such as SRGAN, DenseNet and RCAN, although its Y channel PSNR remains slightly below those of DenseNet and RCAN. This is expected to aid the efficacy of future CADx models, ultimately helping clinicians to accurately identify GI pathologies in WCE in a more confident and timely manner.

## I. Clinical Context and Problem Formulation

Minimally invasive GI diagnostics is highly dependent on wireless capsule endoscopy (WCE) to image depths of the GI tract that are not reachable using push or balloon enteroscopy [1]. A typical swallowable capsule incorporates optical domes, light-emitting-diodes, a complementary metal-oxide-semiconductor or charge-coupled-device image sensor, a power cell, and a radio-frequency telemetry system, enclosed within a cylindrical package of approximately 26 mm in length by 11 mm in diameter. Tens of thousands of images are acquired over hours during the pill’s passage through the GI tract, producing a voluminous corpus for hand-reading or for automated triaging, and each image must be carefully reviewed to identify villous-mucosal villi, vascular smooth muscle, and highly localized mucosal disruptions indicative of Crohn’s disease, GI bleeding, angioectasias, neoplasia, and colonic polyps.

While capsule miniaturization makes this clinically feasible, it necessarily enforces power, thermal, and bandwidth budgets that lead to a trade-off between spatial resolution and temporal sampling. Higher frame-rates mitigates blind spots to propel the capsule through peristaltic contractions, but necessarily come at the cost of lower spatiotemporal resolution, e.g. 336*×*336 frames at 24 fps. Conversely, high pixel count configurations may be low frame rates and miss transient findings or induce blurring between frames. When spatial resolution is coarse, microscopic surface nanobumps, irregularities, and fine microvascular patterning are aliased or blurred, risking missed findings in reading, and diminishing the precision of downstream automated classification/segmentation models.

Software image super-resolution (SR), as a non-invasive post-acquisition solution, can help patients obtain a more resolved anatomy without requiring changes to the capsule hardware or ionising radiation dose. Single-image super-resolution (SISR) can be particularly desirable for WCE streams, as intestinal motility, specular reflections from mucosal mucus and variable illumination make multi-frame registration across successive perspective views more challenging. Yet SISR is an ill-posed inverse problem, as a given low-resolution (LR) observation *I*LR can have many plausible high-resolution (HR) scene solutions. Transformer-based denoising techniques have been previously explored for related biomedical signal quality problems [2].

Mathematically, the degradation process mapping an HR frame to an LR frame at scale factor *s* can be expressed as

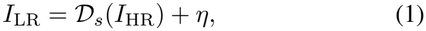

where *D*_*s*_(·) denotes spatial down-sampling (such as bicubic decimation by *s* = 4) and *η* represents sensor and transmission noise. Recovering an estimate *Î*_SR_ requires inverting this many-to-one mapping by leveraging learned structural priors from WCE imagery.

Homogeneous mucosal folds and uniform luminal walls create large low-frequency backgrounds across the frames, while diagnostically-relevant structures – capillary loops, ulcer edges, and pit patterns – exist as localized high-frequency transitions. Standard uniform convolution is applied in both spatial and channel dimensions, which may filter away textures and enhance background noise.

In response to these, we produce a direct solution with our Cascading Feature Refinement with Channel Attention (CFR-CA) for*×* 4 WCE frame super-resolution. Instead of attempting to blindly reconstruct three-channel RGB representations, our entire pipeline decouples the luminance (*Y*) channel from the WCE luminance-chrominance representation in YCbCr, relegating representational capacity to intensity and edge-information, while interpolating a chrominance representation in the frequency domain with two squeezed channels. We did not employ an automated neural architecture search [3] based discovery of the number of blocks *N*, or number of layers in a block *m*, fixed a priori by us.

Our main contributions in this study are fourfold:

- We design the CFR-CA architecture, uniting dense feature cascading with channel-wise attention to preserve low-level mucosal continuity while selectively amplifying high-frequency diagnostic features.
- We integrate adaptive channel re-weighting with compact refinement stages, bottleneck projection, and sub-pixel upsampling to reconstruct sharp *×* 4 super-resolved frames without excessive parameter bloat.
- We establish a YCbCr luminance-centric processing work-flow for WCE super-resolution, demonstrating empirically how targeting the *Y* channel aligns with perceptual sensitivity and improves fidelity metrics.
- We curate a 10,000-image WCE training benchmark stripped of non-informative black capsule borders and benchmark CFR-CA quantitatively, qualitatively, and statistically against established baselines.

## II. Evolution of Deep Super-Resolution in Medical Endoscopy

Neural methods for super resolution from a single image were proposed before the current solutions with end-to-end convolutional mappings significantly outperformed classical Resampling filters such as bicubic or lanczos resampling. Shallow 3 layer convolutional models confirmed the approach by learning the nonlinear patch-hoc mappings from single LR images to HR targets. Deeper models increased the receptive field size by stacking layers for capturing larger spatial support and gradually increased depth as stronger feature hierarchies seemed to allow safer kernel combinations for better edge recovery.

When networks become deeper, difficulty in training and optimization bottlenecks gradually became the main bottleneck against reaching the full potential of the network capacity. For degradation in deep stacks, residual learning solved the problem with deep stacks by letting the layer optimize the residuals with respect to the shortcuts, which are set to the identity mapping; Wider residual structures additionally removed the problems of normalizing too much features and reducing the range of features in SR tasks. However, stacking deep blocks still would lead to diminishing gain, when the early low-level clues were squeezed too much and lost their range in the final fusion/reconstruction blocks.

One way that preserves information is by concatenating the feature maps after each layer in a block, allowing for the direct reuse of low-middle- and high-level features at each subsequent convolution. With a predetermined growth rate, evidence flows forward from the previous convolution, alleviating the vanishing gradient problem while also promoting the sharing of compact features. In our endoscopic frames, the nimbly-shifting mucosal ridges persist with the broad gradient of illumination.

Additional attention mechanisms enhance feature learning by identifying the importance of channels or spatial locations for high-frequency reconstruction. Channel-wise attention estimates inter-channel relationships through global context pooling and gated activation scaling, helping a network suppress non-informative activations while highlighting effective high-frequency filters. Channel attention has been combined with spatial attention for the enhancement of super-resolution (SR) results [4]. Residual-in-residual grouping builds long-range skip connections with channel attention to preserve overall structure while refining details.

Generative adversarial approaches moved SR to perception by adding adversarial networks and perceptual feature losses as detailed above to pixel-wise reconstruction loss. Adversarial and cycle-Consistent losses can generate sharp textures visually similar to ground-truth images. However, hallucination with adversarial losses is a major concern in medical imaging, especially when the added micro-structure of hallucinated regions could be considered as a pathology indicator. Thus, in medical practice, one needs to have a balance of pixel-accuracy, structural similarity, and perceptual naturalness to consider while evaluating the SR approach.

In the scope of endoscopy and general medical imaging, super-resolution methods were used on magnetic resonance image landmarking, diagnostic radiology and conventional endoscopic video enhancement. Existing studies have explored conditional adversarial and attention-enhanced variants of super-resolution on standard endoscopes at different levels of magnification, demonstrating the importance of high-frequency recovery in visual inspection. Super-resolution technology has been assessed for the application on digestive endoscopic images [5]. Yet, customized super-resolution for wireless capsule endoscopy is yet to be fully investigated with specific technical factors in the process, such as the capsule hardware, border artifacts, and the YCbCr luminance domain.

This gap drives us to explore CFR-CA towards the retrieved (expert-chosen) WCE frames. With the integration of dense cascading refinement, the channel-wise attention, and compre-hensive quantitative + statistical evaluation, this work closes the gap between general SR achievement and the actual imaging demands of small-bowel capsule endoscopy.

## III. Proposed CFR-CA Architecture and Mathematical Formulation

The CFR-CA network is engineered to reconstruct high-resolution luminance structure from *×* 4 down-sampled WCE inputs while maintaining stable gradient propagation and strong feature reuse. Given an input luminance patch *X*_0_ ∈ℝ^*H×W ×*1^, an initial 3*×*3 convolutional layer extracts shallow structural features:

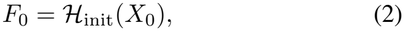

producing 128 feature channels, followed by an initial channel-wise attention stage and a 3*×*3 refinement convolution that conditions early representations before cascading blocks.

After initial feature extraction, the network then passes through a cascade of *N* = 8 sequential CFR-CA blocks (shown vertically in Fig. 1). Each block cascades dense feature build-up through *m* = 8 internal refinement layers with dense feature-wise concatenation, and channel-wise attention and block residual-skip connections. Intra-block fine edges and mucosa folds are learned with local 3*×*3 convolution + rectified linear unit (ReLU) formation; dense concatenation of intermediate feature maps maintains multi-stage contextual cues so that weak edge information is never lost to deep network layers.

**Fig. 1.**
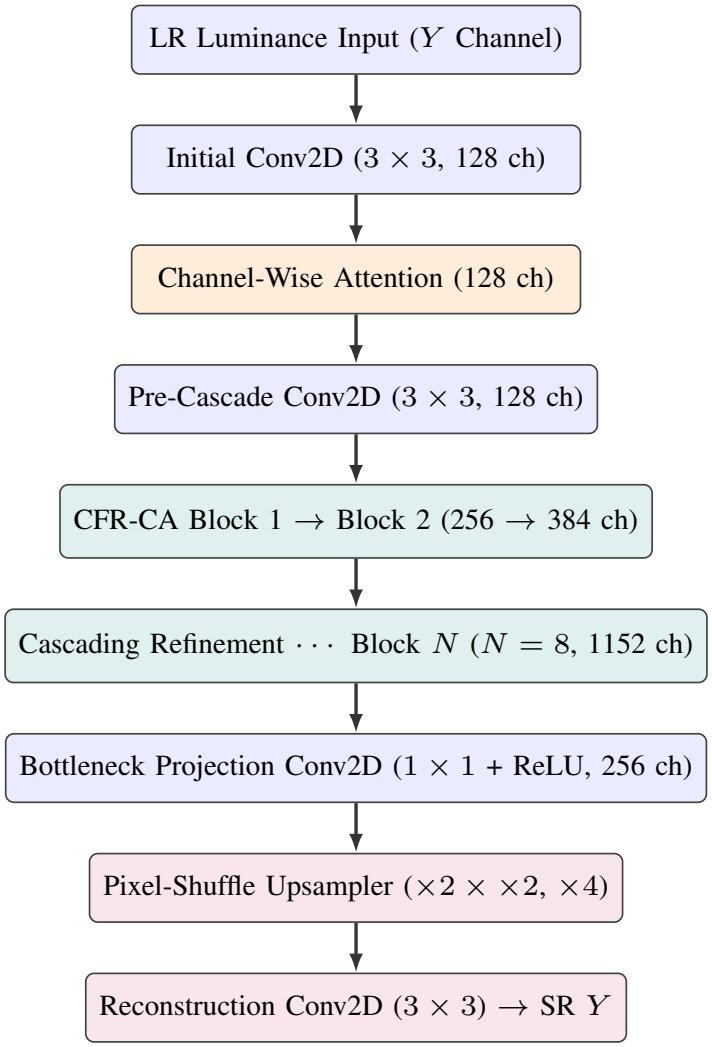
Vertical end-to-end signal flow of the proposed CFR-CA network for *×*4 super-resolution on the WCE luminance channel.

Let *Z*_*l*_ denote the feature representation entering the *l*-th internal layer within a refinement stage. Applying a 3 *×* 3 convolution with ReLU and local residual addition yields

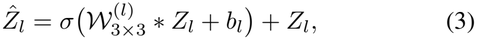

where *σ*(*·*) = max(0, *·*) denotes the ReLU activation, *∗*represents two-dimensional convolution, and 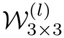 is the learnable kernel. Across cascading stages, feature map widths expand progressively from 128 channels to 256 in the first block, 384 in the second, and up to 1152 across the cascading hierarchy before compression.

To prevent uniform channel weighting from under-representing informative high-frequency filters, CFR-CA incorporates a channel-wise attention module, depicted alongside the cascading refinement block in Fig. 2. Given an intermediate feature tensor *U∈* ℝ^*H×W×C*^ with *C* channels, global spatial context is aggregated via adaptive average pooling:

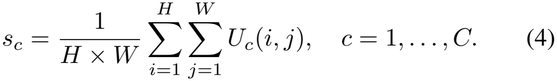

**Fig. 2.**
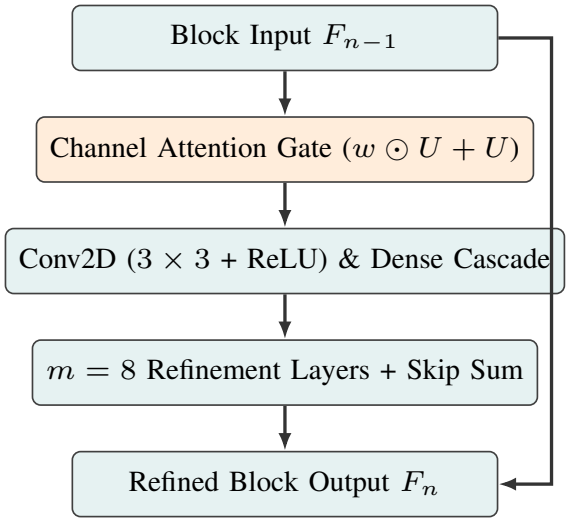
Internal structure of a single CFR-CA refinement block combining channel-wise attention gating, dense cascading refinement, and a block-level residual skip connection.

This channel descriptor is passed through a compact bottleneck transformation using 3 *×* 3 and 1 *×* 1 convolutional layers with ReLU nonlinearity and a sigmoid gating function *δ*(*·*) to produce channel weights *w ∈* (0, 1)^*C*^:

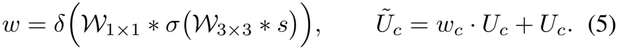

Multiplying each channel by its learned weight adaptively boosts diagnostic texture channels while preserving residual stability through the skip path.

Before spatial upsampling, a bottleneck layer consisting of a 1 *×* 1 convolution with ReLU projects the high-dimensional cascaded representation down to 256 channels. Compressing channel dimensionality at this juncture reduces parameter overhead and computational load during spatial expansion while retaining the most discriminative features learned across the cascading blocks.

Spatial resolution is then restored by a factor of 4 through two sequential sub-pixel shuffling (pixel-shuffle) stages, each providing a *×*2 magnification step:

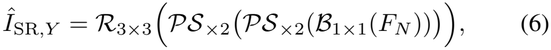

where *B*_1*×*1_ denotes the bottleneck projection, _*×*2_ rear-ranges channel activations into higher spatial resolution grids, and *R*_3*×*3_ is a final 3 *×*3 reconstruction convolution. Operating upsampling in feature space avoids early blur and keeps convolution cost lower than pre-upsampling pipelines.

During training, the network optimizes pixel-level reconstruction accuracy on the *Y* channel using Mean Squared Error (MSE) loss over a batch of *B* training samples:

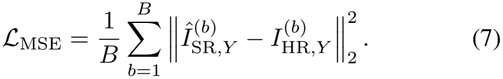

Pixel-wise MSE objectives are a common choice in predictive modelling. At inference time, the reconstructed *Y* channel is recombined with bicubically interpolated *Cb* and *Cr* chrominance channels and converted back to RGB for full-colour visualization, allowing targeted high-frequency refinement without redundant computation on chrominance planes.

## IV. Dataset Curation and Experimental Protocol

We perform super-resolution on real capsule endoscopy data with an image-level training and evaluation set, created from the Kvasir-Capsule public dataset (see Sec. 3.1). This source collection has 47,236 RGB frames taken at 336 *×* 336 pixels from a variety of clinical abnormality and normal appearance categories. Raw frames from the WCE often contain dark peripheral zones and other non-informative areas outside the visible circular or rounded active endoscopic field of view.

We consequently manually inspected and border-cropped the selected frames to remove empty areas that contained only dark background and to ensure the same spatial resolution (280 *×* 280 pixels). This curated dataset comprises 10,000 training, 550 validation, and 1000 test images used to train and evaluate different super-resolution architectures on clean mucosal content.

Paired LR-HR examples are created by using each curated 280*×* 280 image as the reference HR ground-truth image and then down-sampling it with a bicubic factor of *×* 4 to produce the LR input. Every LR input is then converted into the YCbCr colour space, so that network training and inference operates on the luminance (*Y*) channel, which mainly encodes edge contrast and fine structural textures.

Sampled models are uniformly trained on a standard procedure, with the Adam optimizer at a constant learning rate of 0.0001, a batch size of 32, and training time horizon of 300 epochs. The details for data set curation and training hyperparameters are presented in Table I and are shared across the study.

**Table I.**
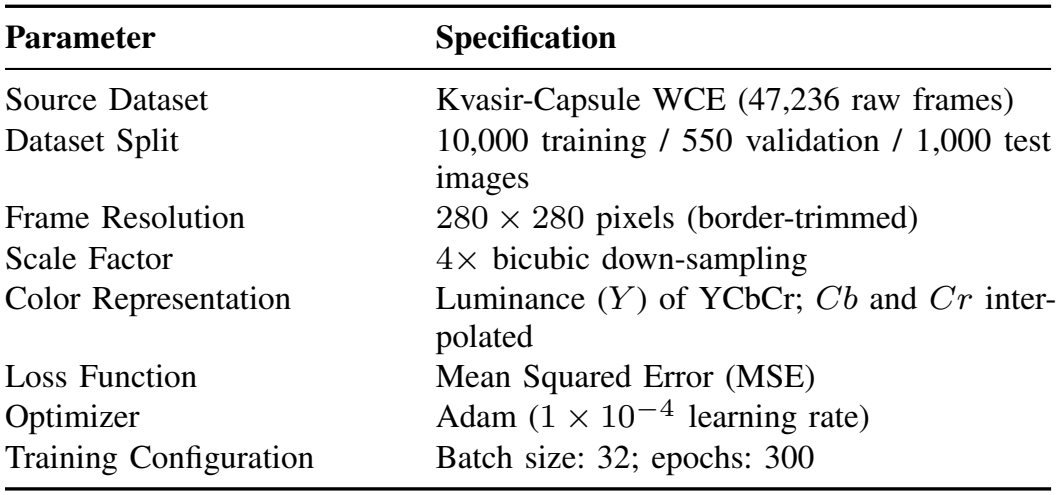
Dataset Curation and Training Configuration.

| **Parameter** | **Specification** |
|---|---|
| Source Dataset | Kvasir-Capsule WCE (47,236 raw frames) |
| Dataset Split | 10,000 training / 550 validation / 1,000 test images |
| Frame Resolution | 280 × 280 pixels (border-trimmed) |
| Scale Factor | 4× bicubic down-sampling |
| Color Representation | Luminance (*Y*) of YCbCr; *Cb* and *Cr* inter-polated |
| Loss Function | Mean Squared Error (MSE) |
| Optimizer | Adam (1 × 10<sup>−4</sup> learning rate) |
| Training Configuration | Batch size: 32; epochs: 300 |

To put CFR-CA in perspective relative to existing approaches, we compare to bicubic plus popular deep SR and image-to-image models such as SRGAN, CycleGAN, Dense Net, and RCAN. By examining classical interpolation and a variety of deep baselines, we gain insight into the benefits of the channel-wise attention and dense cascading strategies.

This can be summarized in terms of complimentary fidelity and perceptual metrics: peak signal to noise ratio (measuring pixel-specific signal fidelity), SSIM (structure, luminance, contrast), and LPIPS (deep perceptual similarity). Lower LPIPS is better.

## V. Quantitative, Perceptual, and Statistical Evaluation

Qualitative comparison of × 4 super-resolved patches and spatial SSIM error maps emphasize the divergences in recovering fine mucosal geometry between competing methods. The loss of continuity in smooth luminal transitions as well as the rounding of fine vascular or villous structures is apparent in many cases as blurring artifacts in SSIM maps. The generative baselines SRGAN and CycleGAN modify the appearance of local contrast; Dense Net and RCAN preserve more structural continuity.

And in all sampled regions, CFR-CA yields tighter edge delineation, and larger high-similarity regions than the bicubic and baseline network outputs. Fine surface ridges and local tissue boundaries are preserved better with less over-smoothing or blockiness.

The quantitative test scores for all the tested methods are listed in Table II. It can be observed that CFR-CA reaches the top of RGB PSNR (40.2261 dB) and is thus better than RCAN, Dense Net, SRGAN, bicubic and CycleGAN (39.4613 dB, 38.8596 dB, 38.0377 dB, 37.2111 dB, 36.9441 dB), respectively.

**TABLE II.** Quantitative Super-Resolution Results (*×* 4) Across Baselines and CFR-CA.

| Method | PSNR (dB) ↑ |  | SSIM ↑ |  | LPIPS ↓ |
|---|---|---|---|---|---|
|  | Y-Ch. | RGB | Y-Ch. | RGB | RGB |
| Bicubic | 38.1069 | 37.2111 | 0.9296 | 0.9057 | 0.2310 |
| SRGAN | 37.0021 | 38.0377 | 0.9291 | 0.9049 | 0.1972 |
| CycleGAN | 38.0121 | 36.9441 | 0.9123 | 0.9012 | 0.1984 |
| DenseNet | 39.6842 | 38.8596 | 0.9401 | 0.9369 | 0.1353 |
| RCAN | 40.1438 | 39.4613 | 0.9427 | 0.9371 | 0.1359 |
| CFR-CA (Proposed) | 39.5389 | **40.2261** | **0.9486** | **0.9378** | **0.1346** |

Also, for the Y channel, CFR-CA obtains 39.5389 dB which is better than bicubic, SRGAN and CycleGAN and close to Dense Net and RCAN (38.1069 dB, 37.0021 dB, 38.0121 dB, 39.6842 dB, 40.1438 dB), respectively.

Structural and perceptual metrics agree the CFR-CA outperforms well as shown in Table II. It achieves the best SSIM (on Y 0.9486, on RGB 0.9378) against RCAN (0.9427 Y / 0.9371 RGB) and Dense Net (0.9401 Y / 0.9369 RGB), and achieves the best LPIPS (0.1346), by a small margin over Dense Net (0.1353), RCAN (0.1359), SRGAN (0.1972), CycleGAN (0.1984), and bicubic (0.2310).

Figure 3 illustrates the mean Y-channel PSNR across models, visually contrasting the jump from bicubic and adversarial baselines to dense and attention-based networks. Alongside mean accuracy, evaluating dispersion across the 1,000 test frames indicates how reliably each method handles diverse GI appearances, lighting gradients, and motion blur.

**Fig. 3.**
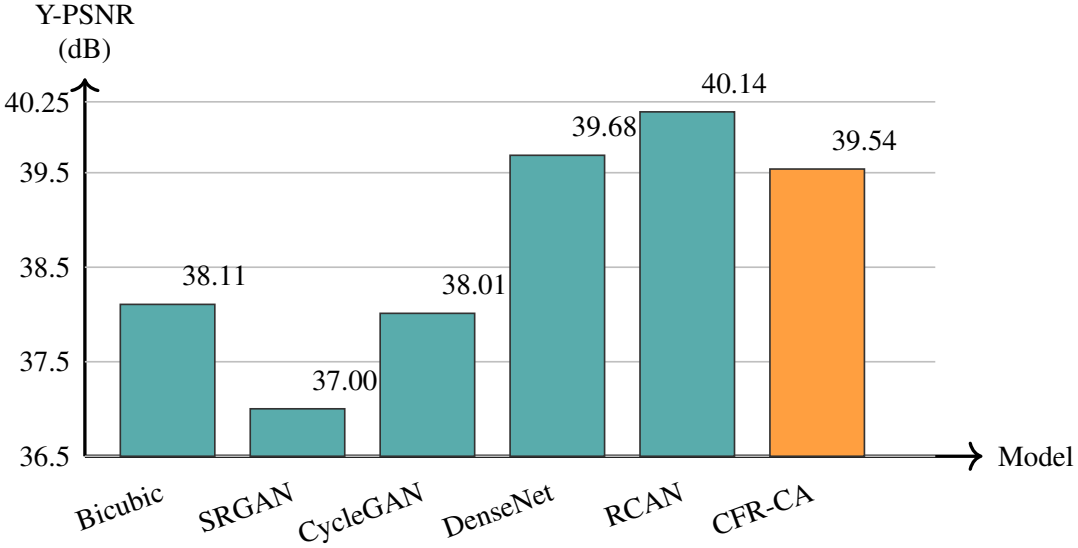
Comparison of mean Y-channel PSNR (dB) across evaluated methods on the WCE test set.

As Table III indicates, CFR-CA has the lowest standard deviation of PSNR for RGB (2.0186) and the lowest standard deviation of SSIM for Y (0.0270) and RGB (0.0306). For the dispersion of Y-channel PSNR, CFR-CA (3.0006) outperforms RCAN (3.5612), SRGAN (3.3490), Dense Net (3.1996), and bicubic (3.8943) with lower Y-PSNR dispersions, while CycleGAN (2.1937) achieves lower Y-PSNR spread at the expense of much larger spread of RGB-PSNR (3.6802) and lower mean accuracy.

**TABLE III.**
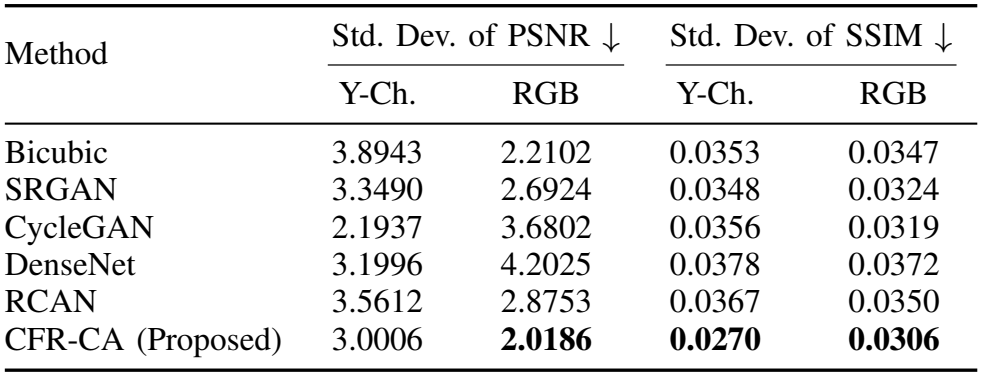
Standard Deviation of PSNR and SSIM Across Test Images (Lower Is More Consistent)

Taken together, the statistical and perceptual results reveal that CFR-CA provides consistently good reconstructions for both smooth intestinal lumen and undulating mucosa during each capsule frame. The consistency in the means shows that there is little fluctuation in SSIM results. This is validated in the low variance of SSIM and illustrates the robustness of CFR-CA.

### VI. Architectural Analysis, Clinical Implications, and Future Outlook

Looking at the CFR-CA architecture, it is clear that the combination of dense feature cascade and channel-wise attention is well-matched for the WCE super-resolution task. The dense connections preserve low-level spatial details directly from the shallow convolutional layers all the way into the later parts of the network. Meanwhile, channel attention acts to amplify high spatial frequency responses over the more uniform tissue backgrounds, and the 1 *×* 1 bottleneck layer prevents the number of channels from growing uncontrollably before the sub-pixel upsampling stage. The high trust clinicians have in this design relates to the interpretability [6].

Further, prioritising computation on the YCbCr luminance plane defines efficiency and metric performance. Fine edges and areas where human visual response is more luminance-dependent than color-dependent mean that enhancing the Y channel enables most structure to be preserved without waste-fully applying the network to the chroma planes. Conversely, because Cb and Cr are only interpolated, a pathology that appears mainly in chroma, with little luminance contrast, cannot benefit from the refinement the network has learned on the luminance channel.

Clinically, however, clearer super-resolved frames could be useful for gastroenterologists scanning long videos, revealing delicate mucosal textures, capillary architecture, and lesion margins. Better structural accuracy of our super-resolution method could also be useful for computer-aided diagnosis algorithms that are trained to recognize signs of disease from capsule images, possibly accelerating early identification of small-bowel disease without the need for hardware upgrades. Computer-aided detection and classification of GI disease from capsule images [7] demonstrates this downstream use.

Some pragmatic limitations should also be taken into account prior to clinical adoption. Training was based on synthetic *×* 4 bicubic down-sampling of centered 280 *×* 280 frames from a single curated derivation, which may not accurately encompass all real-world variations in sensor noise, specular glare, or even dire motion artifacts in transit. Readers should also be aware that quantitative image metrics such as PSNR, SSIM, and LPIPS are no substitute for blinded reading studies that reliably measure clinical diagnostic confidence or pathology detection sensitivity.

Future studies should continue to assess these models on real low-res or degraded capsule frames and explore domain-specific loss functions that couple pixel-perceptual targets with edge or vessel fidelity. Exploiting the synergies of luminance and chrominance upsampling, quantifying end-to-end latency on clinical workstations, and nuanced validation with gastroenterologists will be critical in realizing the potential of super-resolution research in trusted clinical workflows.

## Data Availability

All data used in the present study are from the publicly available Kvasir-Capsule dataset of de-identified capsule endoscopy frames. No new data were collected from patients.

https://datasets.simula.no/kvasir-capsule/

